# Setting up a Hospital Based Diarrhoea Surveillance System in a Low- and Middle-Income Country: Lessons Learned

**DOI:** 10.1101/2024.03.07.24303953

**Authors:** Sam Miti, Caroline C Chisenga, Cynthia Mubanga, Lusungu Msimuko, Chipo Manda, Catherine Zulu, Naomi Muleba Kalaba, Christian Musilikare Niyongabo, Lydia Chisapi, David Thole, Mwizukanji Nachamba, Roy Moono, Moses Chakopo, Dorcas Chibwe, Theresa Kabungo, Kayayi Chibesa, Vivian Nanyangwe, Bwendo Nduna, Gershom Chongwe, Justine Chileshe, Dani Cohen, Roma Chilengi, Seter Siziya, Michelo Simuyandi

## Abstract

**Background:** Acute diarrhoea is a major cause of morbidity and mortality among children in low-resource settings. Establishing effective surveillance systems is crucial for monitoring and responding to diarrhoeal outbreaks.

**Objective:** This manuscript presents the lessons learned during the setup of a hospital-based diarrhoea surveillance system at Arthur Davison Children’s Hospital in Ndola, Zambia. Specifically, the reasons for the delays in processing stool samples from collection to reporting of laboratory results were explored.

**Methods:** The setup of the surveillance system involved several key steps, including stakeholder engagement, training of healthcare workers, development of data collection tools, and establishment of reporting mechanisms. The system aims to capture data on diarrhoea cases admitted to the hospital, including demographic information, clinical presentation and laboratory results.

**Results:** Numerous obstacles were encountered during the implementation of the surveillance. There were three points of delay identified in the ADCH diarrhoea sample handling process from collection to processing: 1) Stool sample collection and packaging 2) Sample transfer from the clinical area to the laboratory 3) Handling and processing in the laboratory. Gaps identified in the three delays related to 1) Staff attitudes and perceptions 2) Health systems infrastructure 3) Operational issues 4) Data management. The following key elements are recommended for setting up a robust, locally owned diarrhoea surveillance system: Implementation of cross-cutting intervention across domains, and a human-centered approach targeted at behavioral change, creating local leadership and ownership of surveillance activities, systematic capacity building through ongoing training/orientation/local data sharing platforms for healthcare personnel, establishing reliable data collection and reporting procedures, addressing infrastructure limitations, and integrating the surveillance system into existing health information systems.

**Conclusion:** We established a hospital-based diarrhoea surveillance system at ADCH in Ndola, Zambia. Several obstacles were identified and resolved, which provide valuable lessons for future implementing of diarrhoea surveillance systems in low resource settings. Successful implementation requires engaging of hospital and laboratory staff, adaptable and easy to use surveillance tools including entering sample information in an electronic laboratory information system and committed leadership.

## Introduction

Globally, acute diarrhoea is the third most common cause of morbidity and mortality in children under 5 years of age (1,2). Despite the successful introduction of rotavirus vaccine in Zambia(3–5), diarrhoea remains a major public health problem with more than 5.4 million episodes reported in under-five children annually (*3*). Of the more than 1,600 public health facilities in Zambia, active acute diarrhoea surveillance is currently ongoing in 3 facilities and only limited to rotavirus. This limitation potentially excludes other enteric pathogens known to contribute to diarrhoea morbidity and mortality in children younger than 5 years of age (6,7). We established a hospital-based diarrhoea surveillance system at the Arthur Davison Children’s Hospital (ADCH) in Ndola, Zambia.

This paper describes the lessons learned during the setup of a hospital-based diarrhoea surveillance system.

## Methodology

### Study Design

An observational study was designed and conducted at ADCH between the 3^rd^ of July 2023 and ended on the 29^th^ of September 2023. ADCH is a tertiary paediatric hospital serving five provinces (Copperbelt, Luapula, Northern, North-Western and parts of Central Province) and has a bed capacity of 250. Additionally, ADCH serves as the first, second and third level health facility for Ndola district which does not have a district or general hospital.

### Human Resource

A multi-disciplinary diarrhoea surveillance team of clinicians, biomedical scientists, nursing, data management, and sample courier staff was established. This was followed by orienting the surveillance team, mapping of key stakeholders within the hospital set-up, reviewing and piloting the data collection tools and SOPs, tracking time from sample collection to processing and overall stool sample workflow.

### Training and Setup

Onsite orientation and training for local clinical and nursing staff, lab teams and sample courier staff on the diarrhoea surveillance system was conducted prior to study roll out. Additionally, staff at ADCH by a team of experts from the Centre for Infectious Disease Research in Zambia (CIDRZ) to strengthen and optimize bacterial culture techniques.

### Stool Sample and Data Collection

Data collection tools were piloted and adjusted accordingly before being rolled out for use. Clinical case-based surveillance forms were completed by the attending clinicians at presentation within OPD and stool samples were collected (where feasible) and sent to the laboratory using a sample courier mechanism.

## Ethical considerations

Ethical approvals obtained from the Tropical Diseases Research Centre Ethics Committe as part of the Shigella Immunology study approval number: TRC/C4/05/2021 Additional permissions were obtained from ADCH to publish this manuscript of lessons learnt during the setup and implementation of a hospital-based diarrhoea surveillance system. The need for consent was waived by the Tropical Diseases Research Centre Ethics Committe because the study was a routine surveillance study. In this study, it was necessary to trace laboratory results back to the patients’ clinical data. As a result, complete anonymity could not be maintained. However, all patient records were stored securely, and access was restricted to only the research staff involved in the study during the study period. This measure was taken to ensure the confidentiality and privacy of the patients’ information while still allowing for the necessary data linkage required for the research.

## Results

We report on lessons learned during the setting up of a hospital-based surveillance system at the ADCH using the 483 participant records for those who managed to submit a stool sample for testing to the lab as shown in Figure 1 below.

**Figure 1:**
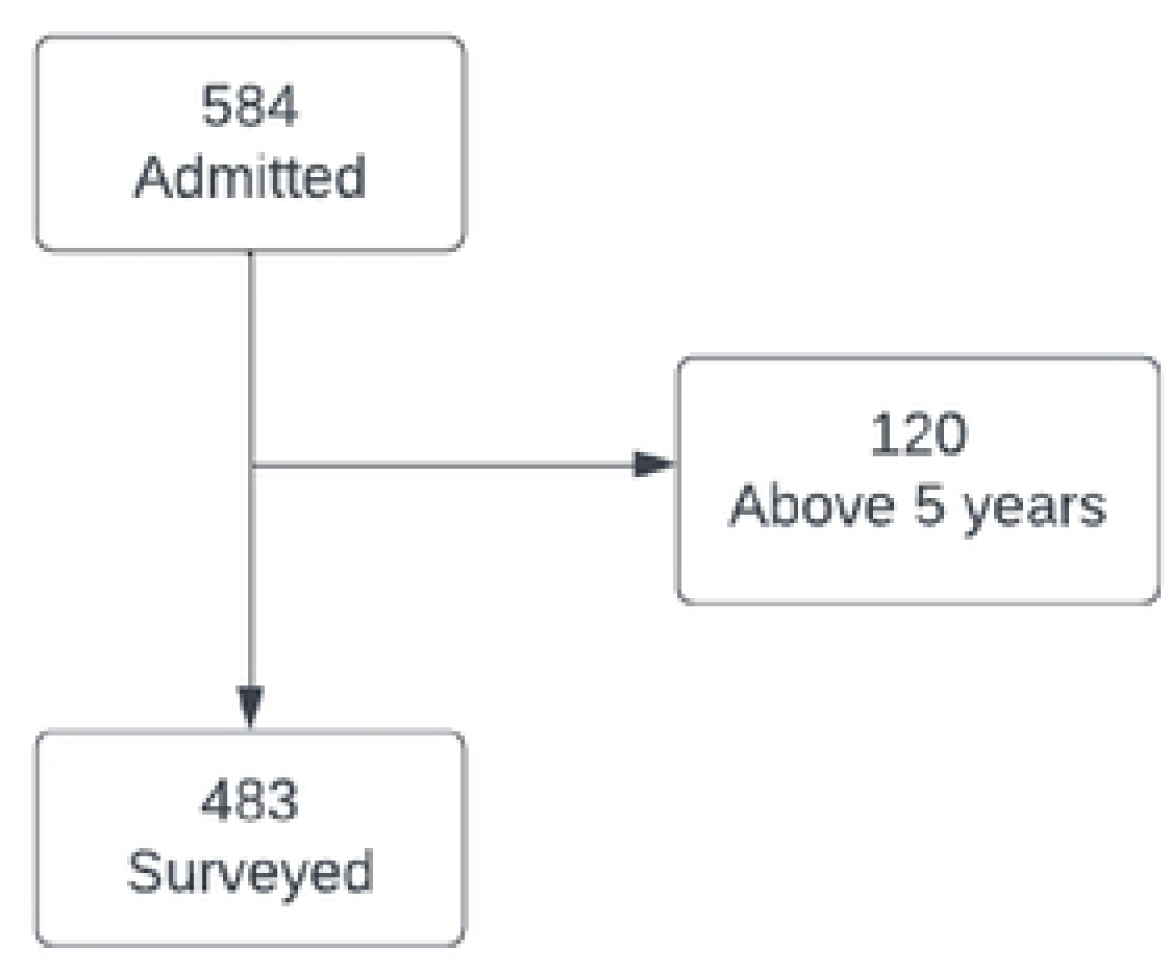
Total number of patients surveyed and those who managed to submit a stool sample.

**Figure 2:**
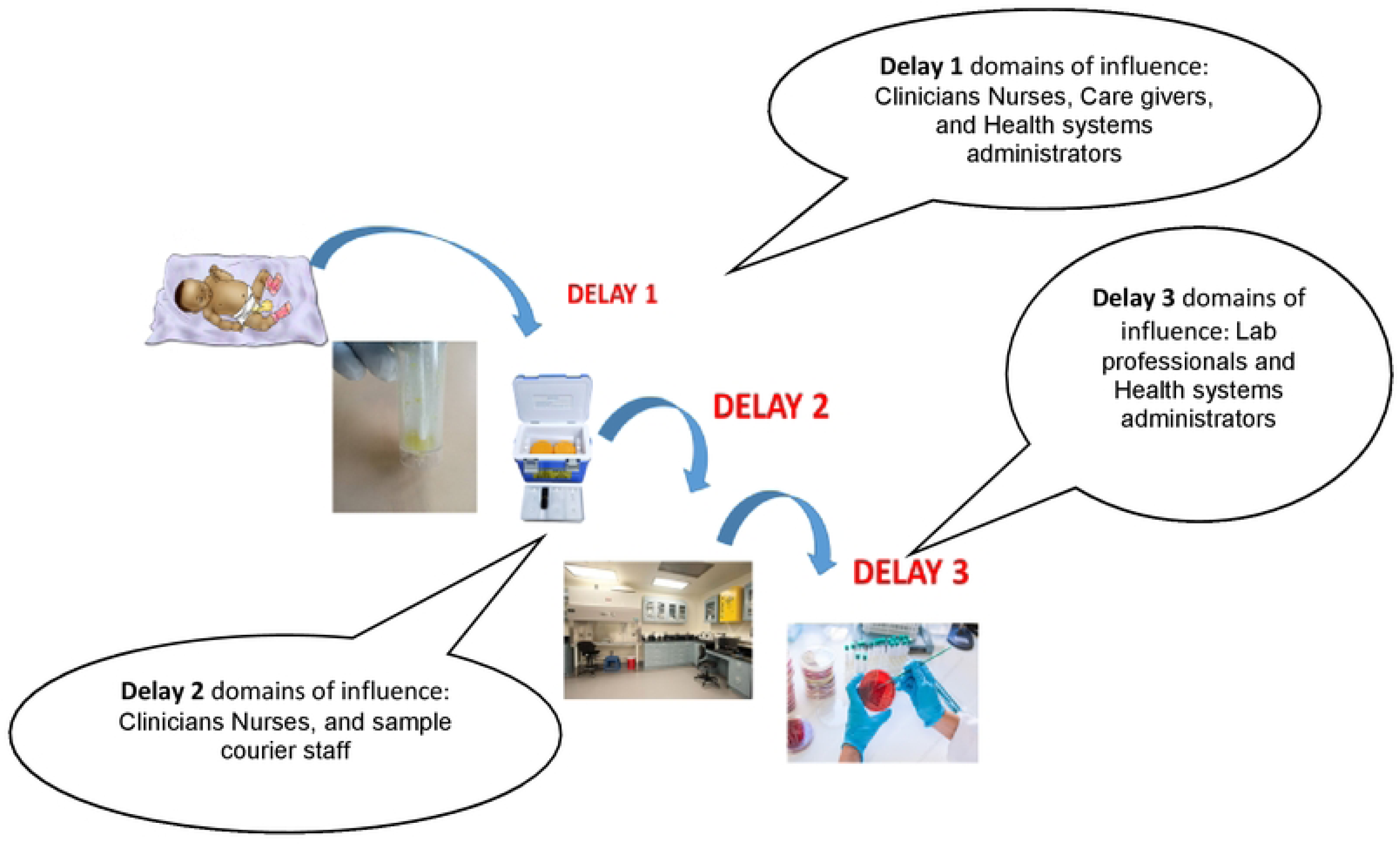
The 3 points of delay in stool sample collection, transport and processing

From the 483 patient files reviewed and surveyed, we noticed 3 main delays from the time of collection to the time of sample processing.

### Delays in stool processing

The three points of delay in handling and processing of stool samples included:

1. 1^st^ Delay-Ccollection and packaging of stool samples: This step involved caregivers coordinating with healthcare providers to timely collect the stool sample and package them for transport to the lab. Examples of activities that were conducted include; failure to collect stool samples from children with watery diarrhea as the diarrhea stool was absorbed by napkin; Health care providers did not clearly communicate with caregivers regarding collected stools leading to disposal of collected samples before submission; health care providers did not adequately inform caregivers on what to do with collected stool.
2. 2^nd^ Delay-Sample courier from clinical area to the lab: Once collected, the stool samples were to be transported from the clinical area by sample courier staff to the laboratory for analysis. However, some delays were observed.in this step such as; stool samples not being considered urgent for immediate transportation to the lab except in combination with other “more urgent ‘’samples like blood samples for pre-transfusion serological analysis, absence of courier sample staff when delegated staff are on leave, paper-based lab information system leading to time wasted on documentation processes.
3. 3^rd^ Delay-Sample handling and processing within the lab: Upon arrival at the laboratory, the stool samples underwent various handling and reception procedures before they could be processed. A significant cause of delay was lack of prioritization of stool samples at the lab reception. For example, stool samples were only considered last after the registry of all other samples for assignment of a unique Lab ID, registry of sample in the Laboratory sample register etc. Once lab reception is completed, technical lab processes include; drying media plates in preparation for culture, transcription of Patient ID and date onto the Culture media plate and entering the sample ID into the microbiology worksheet. Since most reception and lab documentation processes are paper based, they tend to be done in series and not in parallel hence consume a lot of time. This coupled with the lack of direct access by sample courier staff to microbiology section entails that actual stool processing within the lab can only commence once the paper-based documentation processes are complete. This combination of factors contributed to the third delay.

**Table 1:** Delays, their domains of influence, individuals responsible for the delays and recommendations and/or actions that were taken to address them

|  | Description | Identified Gaps | Actions taken |
| --- | --- | --- | --- |
| 1<sup>st</sup><br>Delay | Stool sample collection and packaging | Parents were either afraid /intimidated by staff<br>Parents do not actively look for someone to give the sample | Orientation of clinical and nursing on surveillance and benefits |
|  |  | Parents take long to realize the baby has passed stool | Active engagement of OPD management, shared surveillance findings and involved health systems administrators to find solutions to identified challenges. Administrators supported to use surveillance as ‘Quality improvement process” that enhanced service delivery and improved health worker –care giver communications |
|  |  | Poor attitude by nurses and clinical staff towards the surveillance | Introduced simple and easy interventions that were viewed (stool collections using clean plastics).<br>Creation of demonstration videos, stool collection techniques or innovations<br>Introduction of stool collection SOP using clean plastic |
| 2<sup>nd</sup><br>Delay | Sample courier from clinical area to the laboratory | Poor attitude towards the surveillance | Orientation of courier staff /porter and clinical staff to appreciate role in surveillance efforts |
|  |  | Stool sample not considered urgent and awaits other urgent samples to be shipped | Introduction of stool sample tracking (appendix 1) |
|  |  | Adhering to recommended standard courier | Introduction and task shifting in support of timely courier using nursing student |
|  |  | Patient biological samples transported by hospital staff (porters) in the absence of adequate porter staff | Resources and other staff |
| 3<sup>rd</sup><br>Delay | Stool sample handling and processing within the laboratory | Standard processes demand that sample courier staff (porters) Drop patient samples at Laboratory reception for sample receiving procedures. Stool samples are often not prioritized compared to blood samples for entry into Lab registers and subsequent transportation to microbiology section of laboratory. | Use of stool sample tracking form<br>Active engagement of lab management, shared surveillance findings and involved health systems administrators to find solutions to identified challenges. Administrators supported to use surveillance as 'Quality improvement process' that enhanced service delivery |
|  |  | Poor attitude by Laboratory staff towards stool as a non-urgent sample | Orientation of lab technical and support staff/clerical support staff on diarrhea surveillance and benefits |

**Table 2:**
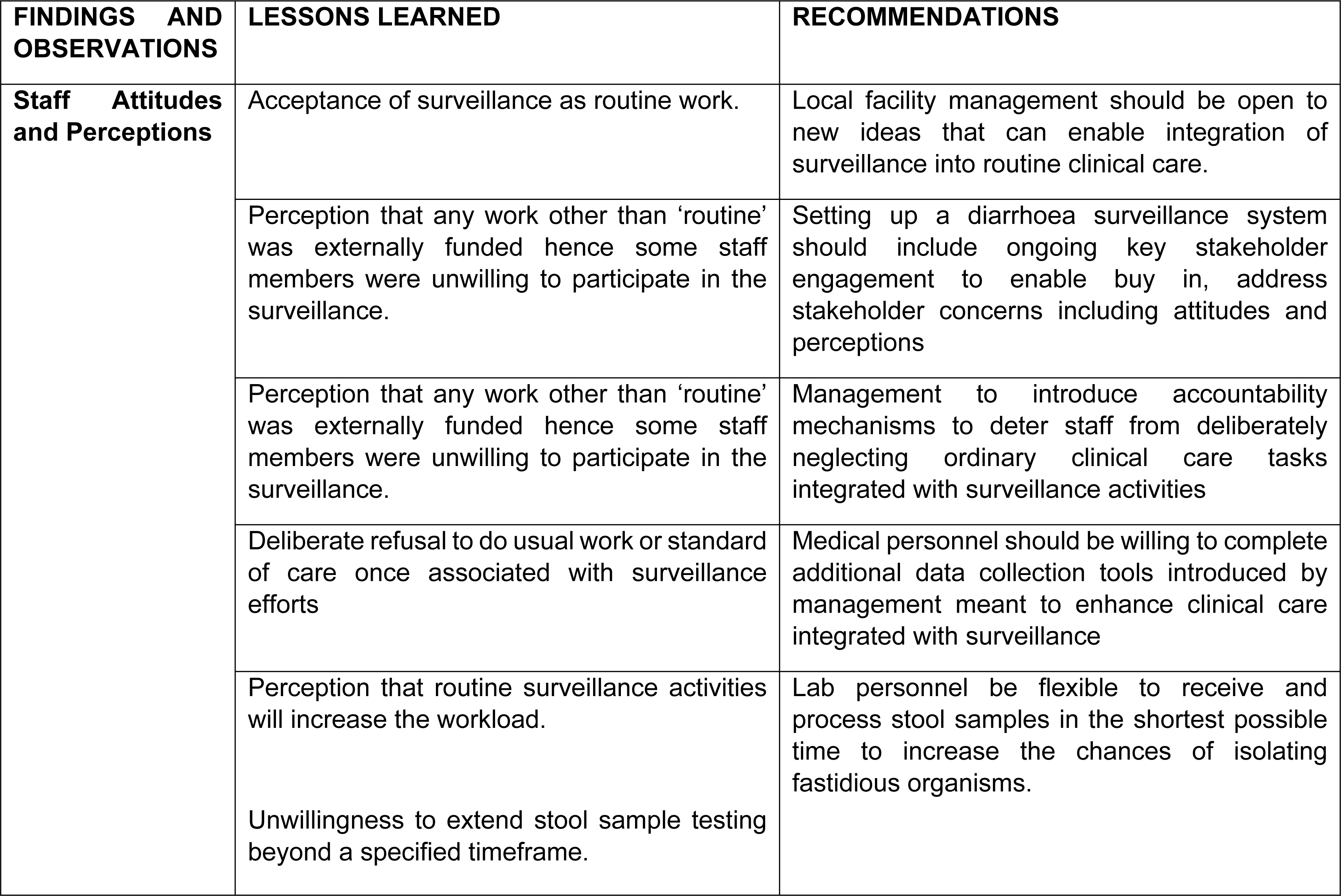
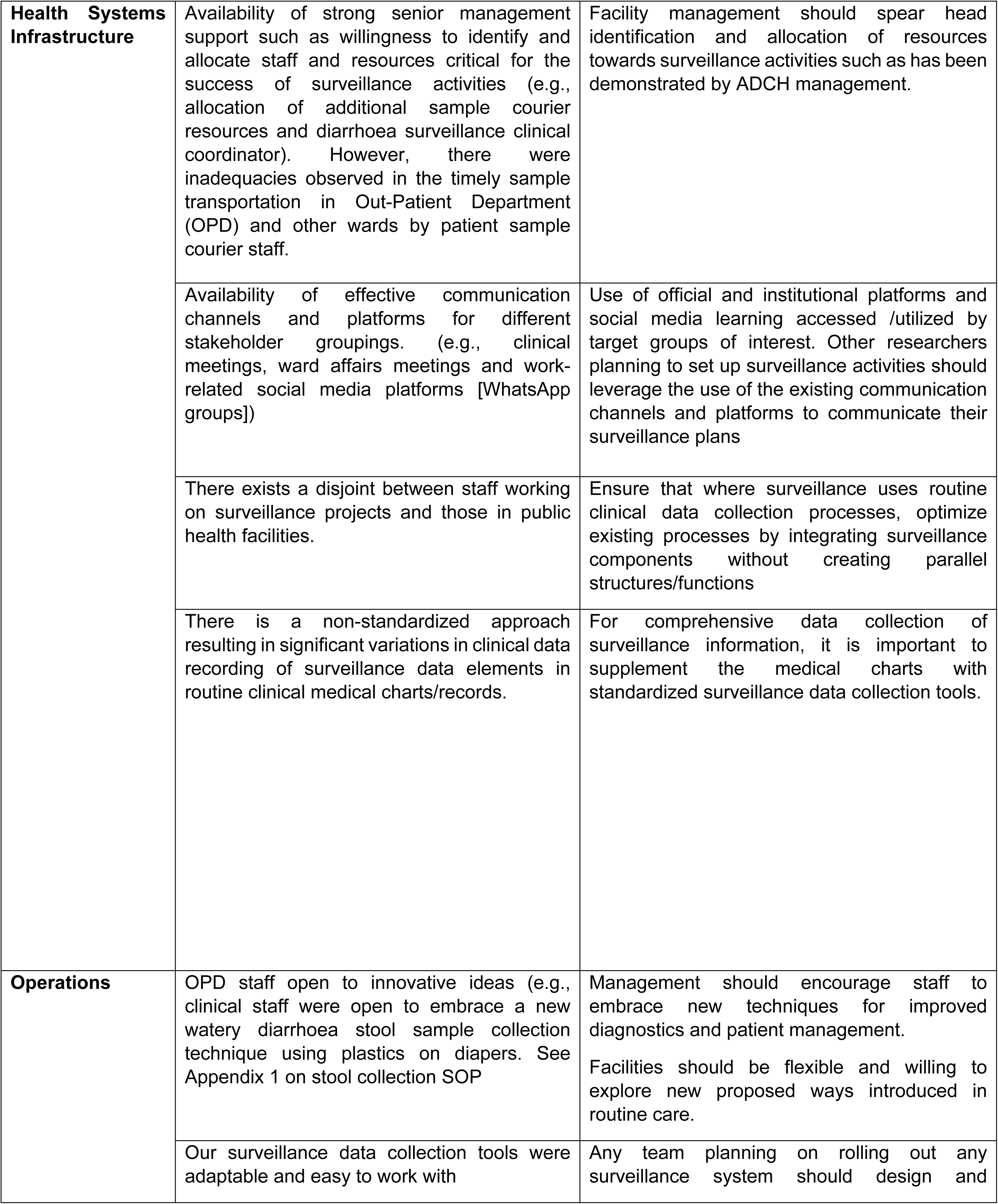
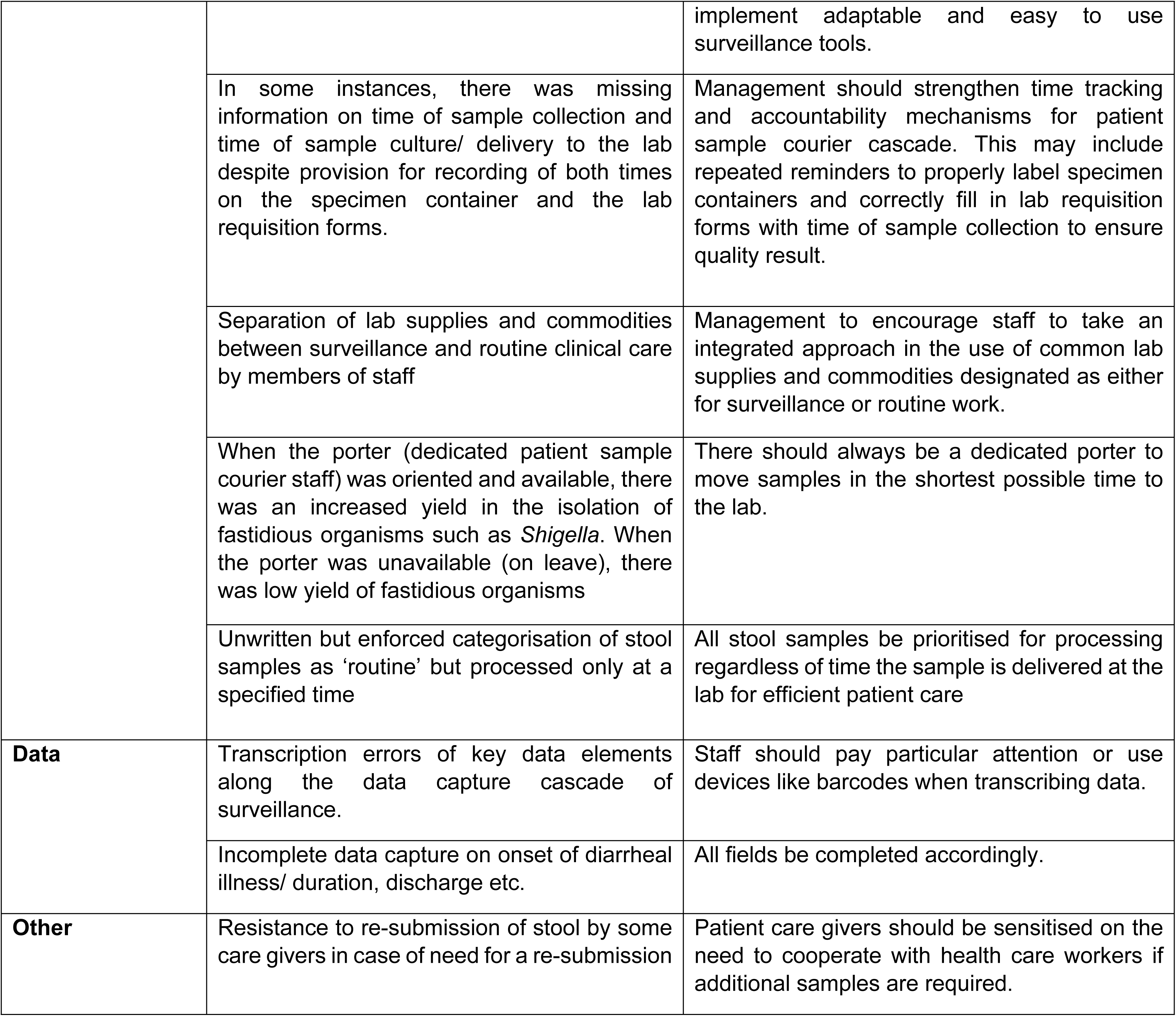
Main challenges, lessons learned and recommendations.

## Discussion

The establishment of a hospital-based diarrhoea surveillance system in Zambia, a low-to middle-income country presented a valuable opportunity to gain insights into the challenges, lessons learned, and recommendations for enhancing the effectiveness of such systems. These observations and experiences can serve as a blueprint for future efforts to strengthen disease surveillance in resource-constrained settings.

Generally, there were three points of delay in handling and processing stool samples observed in our study (1^st^ delay: collection and packaging of stool, 2^nd^ delay: sample courier to lab, 3^rd^ delay: Lab processing of stool). The challenges, lessons learned, and recommendations from the three delays in our study can be broadly categorized into five key areas as (i) Staff Attitudes and Perceptions, (ii) Health Systems Infrastructure, (iii) Operations, (iv) Data, and (v) Other considerations.

### Delays and spheres of influence

Delays in the sample handing and processing cascade have mainly been seen in the context of turnaround time as a whole and not broken down to individual components contributing to the delay(8). Here, we discuss the 3 delays we observed during the set-up of the ADCH Lab based diarrhoea surveillance system:

Our study observed that there were delays in stool collection and packaging in preparation for sample shipment from the clinical area e.g OPD to the microbiology laboratory. These delays involved multiple layers of stakeholders ranging from patient caregivers, immediate healthcare providers to health systems administrators. Unfortunately, most literature on diarrhea management or treatment lacks focus on the collection, barriers to collection, packaging, and timely transportation to the lab (9), with discussions primarily centered on general diarrhea management, etiology such as Cholera and Shigellosis/dysentery (10) (11), or more recently on Antimicrobial resistance (AMR) (12) (13). Regrettably, diarrhea stool collection or handling process-as central to etiologic-pathogen isolation, are often not exhaustively discussed. Delays in reporting laboratory results can have a significant impact on patient diagnosis and treatment decisions (15) (16) as factors related to microbe stability, such as time and temperature, are major contributors to false results (17) especially for fastidious organisms such as *Shigella*. For example, Shigella-positive stool samples stored at room temperature for more than 45 minutes, may be culture negative unless stored at 4 degrees or less (18). There is paucity of data that specifically addresses delays in the context of clinical specimen collection and handling prior to lab transportation, other samples other than HIV, COVID 19 or blood samples (18) (19) (14). Stool sample collections or diarrhoea samples are rarely discussed in the context of individual delays in the sample collection from patient to lab processing cascade (16). Delayed sample collection and processing can have significant implications for antimicrobial resistance (AMR) in low- and middle-income countries (LMICs). If stool samples or diarrhoea samples are not collected and processed in a timely manner, there is a risk that bacterial infections may go undetected. This can lead to empirical treatment with broad-spectrum antibiotics, which may not be effective against the specific bacteria causing the infection. Overuse of antibiotics due to delayed or missed diagnoses can contribute to the development of AMR, as bacteria become resistant to commonly used antibiotics (20) (21). Therefore, addressing delays in sample collection and processing is crucial for controlling the spread of AMR in LMICs. Efforts to improve laboratory infrastructure, training of healthcare workers, and public health education on the importance of timely sample collection are essential in combating AMR in these settings.

The second area of delay was transferring the samples by courier from the clinical area to the laboratory. Health worker attitudes, a limited number of courier staff and courier processes were observed as responsible for delays in transferring stool sample to the lab in a timely manner. Health workers generally did not consider stool samples as urgent to be transported alone unless in combination with other “more urgent samples” such as blood or CSF. This action significantly contributed to the stool samples being kept in unfavorable temperature conditions for longer periods and possibly negatively affected the isolation or detection of enteric pathogens such as *Shigella*. For example, Miti et al 2023 estimated that 99% of Shigellosis in Zambia could be missed based on extrapolation of DNA PCR testing of stools and HMIS reported cases of dysentery (22). Thus, the low reported cases of Shigella/dysentery could have been due to the insensitivity of using clinical diagnosis of dysentery and poor culture yields of *Shigella* even when PCR results were positive (23). Our findings of low reported cases of Shigella/ poor culture yields even when PCR results were positive are consistent with findings from other published studies (24) (19). Nurses, clinicians, sample courier staff, and health system administrators are identified as key stakeholders who can positively influence this stage by creating favorable conditions for prompt courier of stools.

The third area of delay was the handling and processing of stool samples within the laboratory. This study observed that standard operating processes require that sample courier staff (porters) drop patient samples at Laboratory reception for sample receiving procedures. This coupled with poor attitude by some laboratory staff towards stool as a non-urgent sample causes stool samples to sit at the lab reception for prolonged periods. Efficiently entering patient details into a system or registers upon sample arrival at the laboratory is highlighted as important for accurate patient identification and tracking throughout the testing process (25). However, if the documentation is solely paper based, such documentation processes add to significant delay in sample processing. We recommend the introduction of barcoding of all samples so that once any sample is received at the lab, it can only be scanned and all details would then be obtained and verified in order to reduce the delay in sample processing (26) (27) Implementing an electronic Laboratory Information System (LIS) is recommended as a time-saving measure for patient data entry (24). Laboratory professional staff and administrators, as well as health system administrations, are identified as stakeholders who can positively influence this stage through appropriate health system infrastructure such as barcode scanners etc.

To establish successful hospital-based diarrhea surveillance, it is crucial to address delays at various stages of the surveillance cascade. This requires a thorough understanding of potential sources of delay and their causes in a specific context. Interventions should be designed to target stakeholders responsible for specific delays in the handling of stool samples. It is also important to engage relevant stakeholders and consider biological determinants of microbe stability during collection, storage, transportation, and processing within the laboratory when designing interventions (28) (29) (30).

### Staff Attitudes and Perceptions

One of the fundamental aspects influencing the success of our surveillance system was the attitudes and perceptions of healthcare staff involved. Resistance to change and a lack of awareness about the importance of surveillance systems posed initial challenges. However, as the system became operational, staff members increasingly recognized its significance in timely disease detection and response. This highlights the need for comprehensive training and continuous engagement to foster a positive attitude towards surveillance activities (31) (32). Our results align with those of a study carried out in India (33). In that study, after the establishment of materials and a communication system, a sequential training program was devised. This program encompassed training at the district, block, and community levels. It involved instructing trainers from each block and conducting multiple training sessions for community volunteers at the village level. Importantly, this project was integrated into the pre-existing surveillance and reporting framework.

### Health Systems Infrastructure

The adequacy of health systems infrastructure played a pivotal role in determining the functionality of our surveillance system. Inadequate resources, including laboratory diagnostic supplies and consumables and an unreliable supply chain, resulted in delays in data collection and analysis. Our observations align with the findings of Melania et al., who discovered that laboratory capacity frequently represents the primary constraint in laboratory-based surveillance systems (34). Our experience underscores the imperative of investing in robust health infrastructure to support surveillance initiatives. Furthermore, integrating surveillance activities into existing healthcare structures can optimize resource utilization and sustainability.

In addition, strong senior management support, including a willingness to identify and allocate critical staff and resources, proved essential for the success of surveillance activities. For instance, the allocation of additional sample courier resources and the appointment of a diarrhoea surveillance clinical coordinator were key components. However, challenges were observed in the timely transportation of samples from the Out-Patient Department (OPD) and other wards by patient sample courier staff. As noted in a research conducted by Jayatilleke and colleagues, it is crucial to delineate the roles and responsibilities of each individual involved in the surveillance system to ensure its seamless operation (35).Trained staff in adequate numbers are essential for this purpose.

Effective communication channels and platforms for various stakeholder groups, such as clinical meetings, ward affairs meetings, and work-related social media platforms (WhatsApp groups), were instrumental in facilitating coordination and information exchange. However, a significant gap was noted between staff engaged in surveillance projects and those in public health facilities, highlighting the need for greater integration to enhance clinical care and patient management. In accordance with Malania et al.’s research, they underscored the critical role of strong diagnostic stewardship at every phase of diagnostic practice for the effectiveness of a surveillance program. This stewardship covers guidelines for specimen selection and collection, the handling of clinical samples in the laboratory, and the reporting and analysis of results. Effective communication among all stakeholders is essential for these elements to function cohesively.

### Operations

Operational challenges encompassed the day-to-day management of the surveillance system. These included issues related to staffing, logistics, and coordination among different healthcare units. According to the study by Lim and colleagues, establishing a strong and committed connection between the proficient microbiology laboratory and healthcare practitioners is vital. This relationship not only enhances mutual confidence but also maximizes the effective utilization of available diagnostic resources (36).

Ensuring a dedicated team, streamlined logistics, and clear communication channels are pivotal for the smooth operation of surveillance systems. Consistent with the study conducted by Malania et al., their research emphasized the importance of a multifaceted or multidisciplinary approach that hinges on a functional infectious disease diagnostic cycle. This cycle encompasses clinicians collecting and submitting clinical samples to a microbiology laboratory, a bacteriology laboratory equipped for species identification and antimicrobial susceptibility testing (AST), as well as a well-structured system for reporting, consolidating, analysing, and interpreting data to guide actionable decisions (34).

### Data Management

Effective data management emerged as a cornerstone of our surveillance system. Challenges included data quality, timeliness, and security. Lessons learned emphasize the importance of standardized data collection tools, continuous data quality checks, and secure storage and transmission of data. Furthermore, the integration of digital technologies can enhance data management efficiency and accuracy. Our study corroborated the findings reported by a south African study, where some of the important lessons that the technical staff learned in setting up a surveillance system included designing good data collection instruments, database set up and data management (37).

### Other Considerations

In addition to the above categories, other considerations such as community engagement, public awareness, and resource mobilization played significant roles in our surveillance system’s success. Community involvement in reporting diarrhoea cases and raising awareness about the importance of early reporting contributed to enhanced case detection. Moreover, securing sustainable funding sources and fostering partnerships with local and international organizations were instrumental in maintaining the system’s functionality.

The hospital-based surveillance system was successfully rolled out and various key stakeholders were engaged and trained on the data collection tools.

There were various strengths observed in this study; Firstly, the study resulted in improved capacity for lab, clinical and data personnel, and lab staff become more alert to other enteric pathogens present in the stool.

Secondly, there was improved time-to-processing, which led to quality results and improved patient management. Thirdly, we observed increased awareness among medical personnel with regards to the burden of diarrhoea disease, the emerging pathogenic organisms, and the need for timely submission of stool samples to the lab. Fourthly, this study contributed to strengthened isolation and storage capacity of fastidious organisms such as *Shigella*. Finally, enhanced collaborative activities among Centre for Infectious Disease Research in Zambia (CIDRZ), Centers for Disease Control and Prevention (CDC), Tropical Diseases Research Centre (TDRC) and Arthur Davison Children’s Hospital (ADCH).

Despite the notable strengths, there were some challenges encountered, broadly grouped as; *Clinical* (A few clinicians were not making use of the forms, a good number of lab forms had missing information on time of collection, inconsistent use of plastics for collection and not all patients surveyed submitted a stool sample.)

*Lab* (lab staff were unable to conduct Antimicrobial Sensitivity Testing on *E-coli* strains, discarded stool samples before storage, did not indicate time of culture, inefficiencies in moving samples from lab reception into microbiology section.)

*Data* (mismatching patient identification numbers, missing of key participant information on the data case)

*Other* (staff attitude such as being slow to change despite conducting training and sensitisation, not willing to receive samples beyond specified times and unwillingness to complete the data collection tools).

## Conclusion

Diarrhoea is still a problem in Zambia and there is need to extend the current established surveillance system at ADCH to other facilities in order to understand the burden and the circulating pathogens comprehensively. Setting up a hospital-based diarrhoea surveillance system in a low-to middle-income country is a complex endeavor, but one that can yield substantial public health benefits. The challenges and lessons learned from our experience underscore the need for a holistic approach that addresses staff attitudes, health systems infrastructure, operational efficiency, robust data management, and other considerations.

## Recommendations

We therefore recommend that going forward, there is need (i) to establish hospital-based diarrhoea surveillance systems in other facilities, (ii) for continued staff sensitisation and training with regards to the burden of diarrhoea in under-five participants, (iii) for dedicated resource to test for *E-coli* to understand the distribution of the pathogenic strains and their antimicrobial resistance profiles. The following are our recommendations:

1. Establishing Diarrhoea Surveillance Systems: We recommend the establishment of hospital-based diarrhoea surveillance systems in other facilities to enhance disease monitoring, preparedness and response capabilities.
2. Staff Sensitization and Training: Continued staff sensitization and training regarding the prevalence and impact of diarrhoea in under-five participants is essential for effective surveillance.
3. Resource Allocation for *E-coli* Testing: Dedicate resources for *E-coli* testing to better understand the distribution of pathogenic strains and improve targeted interventions.
4. Capacity Building: Invest in ongoing training and capacity-building programs to cultivate positive staff attitudes and enhance surveillance skills.
5. Infrastructure Enhancement: Prioritize investments in healthcare infrastructure to ensure the availability of necessary resources for timely and accurate data collection and analysis.
6. Streamlined Operations: Establish efficient operational protocols, including clear staffing structures, logistics management, and interdepartmental coordination.
7. Improved Data Management: Implement standardized data collection tools, stringent data quality checks, and secure digital data management systems to enhance data accuracy and accessibility.
8. Community Engagement: Engage with local communities to encourage early reporting of diarrhoea cases and raise awareness of the importance of diarrhoea surveillance.
9. Resource Mobilization: Seek sustainable funding sources and establish partnerships with relevant stakeholders to secure ongoing support for surveillance activities.
10. Utilization of Available Resources: Make efficient use of existing resources by strategically identifying and allocating them to support surveillance activities.
11. Communication Channel Utilization: Effectively utilize existing communication channels and platforms to facilitate collaboration and communication regarding surveillance plans.
12. Promote Integration: Encourage integration and collaboration between staff involved in surveillance projects and those working within public health facilities to enhance clinical care and patient management.
13. Standardize Data Collection: Ensure comprehensive data collection of surveillance information by incorporating standardized surveillance data collection tools into medical charts.
14. Barcode Implementation: Consider implementing measures such as barcodes or similar devices to enhance the accuracy of data transcription in the surveillance system.
15. Comprehensive Data Completion: Ensure that all fields in the surveillance data collection process are completed accurately and comprehensively to maintain data integrity and reliability.
16. Patient Caregiver Sensitization: Sensitize patient caregivers to the importance of collaborating with healthcare workers in cases where additional samples are required for the surveillance system.

By adhering to these recommendations, future efforts to establish and maintain hospital-based diarrhoea surveillance systems in low-to middle-income countries can address challenges, enhance effectiveness, and contribute to improved patient management and public health outcomes. Our research resonated with the conclusions reached in a study carried out in Georgia, which emphasized that the successful implementation of a laboratory-based surveillance system in low- and middle-income countries is contingent upon its alignment with the country’s existing capacity levels (34)

## Data Availability

All relevant data are within the manuscript and its Supporting Information files.

## ACKNOWLEDGMENTS

We would like to express our gratitude to all the individuals and organizations who have contributed to the development and completion of this manuscript titled “Setting up a Hospital Based Diarrhoea Surveillance System in a Low- and Middle-Income Country: Lessons Learned.”

Primarily, we would like to thank the staff and management of the Arthur Davison Children’s Hospital for their cooperation and support throughout the project. Their dedication and commitment to improving healthcare in their communities have been invaluable.

We are also grateful to the Clinical team as well as the Microbiology Lab professionals at Arthur Davison Children’s Hospital who participated in the surveillance system and provided valuable insights and support. Their expertise and willingness to share their experiences have been crucial in shaping the lessons learned that are presented in this manuscript.

Furthermore, we would like to acknowledge the enteric disease unit of the Centre for Infectious Disease Control (CIDRZ) for their support, without which this work would not have been possible. Their investment in public health research is essential for addressing healthcare challenges in low- and middle-income countries.

Finally, we extend our thanks to the reviewers and editors at PLOSONE journal for their time, effort, and constructive feedback on this manuscript. Their input has undoubtedly improved the quality of our work.

We are sincerely grateful for all the support we have received, and we hope that this manuscript will contribute to advancing healthcare systems in low- and middle-income countries.

## Notes

### Competing Interest Statement

The authors have declared no competing interest.

### Funding Statement

The author(s) received no specific funding for this work.

### Author Declarations

The Tropical Diseases Centre Ethics Committee granted approval for this study (study approval number TDRC/C4/05/2021)

